# Altered Reward-Related Resting-State Network Properties in Adolescent Cannabis Use and Depression

**DOI:** 10.1101/2025.11.01.25339294

**Authors:** Tram N. B. Nguyen, Russell H. Tobe, Benjamin A. Ely, Vilma Gabbay

**Affiliations:** Department of Psychiatry and Behavioral Sciences, Albert Einstein College of Medicine, Bronx, NY, USA; Medical Scientist Training Program, Albert Einstein College of Medicine, Bronx, NY, USA; Nathan S. Kline Institute for Psychiatric Research, Orangeburg, NY, USA; Department of Psychiatry and Behavioral Sciences, University of Miami Miller School of Medicine, Miami, FL, USA; Center for the Developing Brain, Child Mind Institute, New York, NY, USA; Department of Psychiatry, New York University Grossman School of Medicine, New York, NY, USA

**Keywords:** fMRI, Graph Theory, Reward, Adolescent, Cannabis, Depression

## Abstract

**Objective:** Cannabis use among adolescents with depression is prevalent. Reward dysfunction has been documented in each condition separately, yet rarely examined in their co-occurrence. Here, we investigated cannabis use and depression comorbidity in relation to resting-state properties of reward networks in a trans-diagnostic adolescent sample.

**Method:** Clinicians interviewed adolescents and assessed depression with the Children’s Depression Rating Scale. Cannabis use was characterized by self-report and toxicology screens. Neuroimaging scans were acquired and processed using the Human Connectome Project pipelines. Participant-level resting state data were parcellated, and *Reward Expectancy* and *Reward Attainment* network masks derived from a reward task were applied. Graph theoretical metrics, including Strength Centrality (C_Str_), Eigenvector Centrality (C_Eig_), and Local Efficiency (E_Loc_), were estimated within each network. Group-level clinical correlates of network properties were assessed with non-parametric analyses (10,000 permutations), adjusted for age, sex, and family-wise error (FWE) rates (p_FWE_ < 0.05).

**Results:** In the full sample (N = 131; 15.3 ± 2.2 years; 65.2% female), depression severity was associated with stronger E_Loc_ of the ventral striatum within the *Reward Attainment* network. Among those who used cannabis (n = 38), heavier cannabis use was linked to weaker C_Str_ of the anterior cingulate cortex and E_Loc_ of the postcentral somatosensory area within the *Reward Expectancy* network. Exploratory analyses further revealed cannabis-related dorsolateral prefrontal and cerebellar dysconnectivity across the whole brain, as well as sex differences.

**Conclusions:** Our findings suggest that adolescent cannabis use and depression differentially influence resting-state reward network properties in co-occurring context. Additional research with larger cohort sizes is needed to corroborate findings.

## 1. Introduction

Adolescent cannabis use is prevalent, with 34% of 12^th^ graders in the United States having tried cannabis and 13% reporting daily use for at least a month^1^. Alarmingly, adolescents with depression are more than twice as likely to use cannabis as their peers without depression^2^ and face heightened risk for disordered use^3,4^. During adolescence, the endocannabinoid system plays a critical role in the maturation of neural circuits^5^, and disruptions of endocannabinoid modulation can potentiate depressive and substance use disorders^6,7^. Animal studies show that Δ^9^-tetrahydrocannabinol (THC), the primary psychoactive agent in cannabis, disrupts the adolescent reward system and induces depressive-like behaviors^5,8–10^. Consistent with these preclinical findings, youth who use cannabis exhibit motivational deficits^11^, including higher levels of anticipatory anhedonia^12^, reflecting diminished capacity to expect pleasurable rewards (i.e. reward expectancy). Neuroimaging studies in youth who use cannabis have also implicated alterations in reward circuitry,^13–15^ a system central to the development and maintenance of depression^16^. Together, these data suggest that reward dysfunction may represent a shared mechanism underlying the co-occurrence of adolescent cannabis use and depression. However, direct neuroimaging investigation of their co-influence on the developing reward system is limited, as studies on cannabis use rarely account for depression, while those on depression typically exclude adolescents who use cannabis.

Resting-state functional magnetic resonance imaging (rs-fMRI) is a powerful modality for studying functional brain networks without task engagement. In rs-fMRI, the most common analytic approach is seed-based functional connectivity. In the few existing studies on adolescent cannabis use, this method has revealed connectivity alterations in frontal, parietal/sensorimotor, and cingulate cortices^17^. Concurrently, impaired functional connectivity centered on cingulate, limbic, and striatal regions has been reported in adolescent depression^16,18,19^. While seed-based analyses have identified large-scale network dysconnectivity in each condition, they have provided only modest insight into the integrity of networks fundamental to reward processes. Addressing this methodological limitation, our group has implemented the fMRI Reward Flanker Task (RFT) to study neural processing during distinct phases of *Reward Expectancy* and *Reward Attainment*^20,21^. We then extracted network masks from the most strongly recruited regions in each reward phase and applied these reward network masks to resting-state data, employing graph theory to identify nodes where altered network topography was associated with depression and anxiety symptomatology^22^. Particularly, we detected significant associations between depression severity and altered striatal communication within the *Reward Attainment* network^22^. Graph theoretical approaches provide a framework to comprehensively characterize network organization and identify key information processing hubs^23^. Importantly, as graph theoretical approaches are data-driven, they offer a robust description of connectivity during sensitive developmental periods^24^ and can reveal subtle anomalies not captured by seed-based methods.

Capitalizing on this innovative approach, here we aimed to investigate resting-state properties of *Reward Expectancy* and *Reward Attainment* networks in relation to cannabis use and depression in adolescents. Based on prior findings, we hypothesized that: 1) Cannabis use would be associated with altered properties of the *Reward Expectancy* network; and 2) Depression severity would be associated with altered properties of the *Reward Attainment* network. Given well-established sex effects in cannabis use^15^ and depression^25^, we also conducted exploratory sex-stratified analyses. Exploratory whole-brain analyses were also performed to assess neural organizations beyond *a priori* reward networks.

## 2. Methods

### 2.1. Participant Recruitment and Initial Assessments

In our research program focusing on depression and related conditions, we recruited youth from the New York metropolitan region through clinician referrals and community-based advertisements. Study procedures were approved by the Institutional Review Boards (IRB) of the University of Miami Miller School of Medicine, Nathan Kline Institute for Psychiatric Research, Albert Einstein College of Medicine, and Icahn School of Medicine at Mount Sinai. General eligibility was assessed in initial phone interviews. For adolescents younger than 18 years, assent was obtained, and a parent/guardian signed informed consent. Adolescents 18 years and older provided their own informed consent.

During the clinical assessment visit, a trained child and adolescent psychiatrist or a licensed clinical psychologist conducted semi-structured diagnostic interviews to determine psychiatric symptomatology and DSM-5^26^ diagnoses for each participant. Participants younger than 18 years were evaluated with the Kiddie Schedule for Affective Disorders and Schizophrenia – Present and Lifetime Version for Children (K-SADS-PL)^27^, while those aged 18 years and older underwent the Mini International Neuropsychiatric Interview (MINI)^28^. On the scan day, fasting blood draws were performed to assess complete blood count, thyroid function, and liver function. A urine toxicology test was also administered to screen for amphetamines, barbiturates, benzodiazepines, cocaine, 3,4-methylenedioxymethamphetamine (MDMA), methamphetamine, opiates, oxycodone, propoxyphene, and THC. Female participants completed urine pregnancy tests.

### 2.2. Inclusion and Exclusion Criteria

Eligible participants were 12-21 years old, of any sex, race, or ethnicity. Following a dimensional approach, participants with any level of cannabis use or depression were included, regardless of whether they met DSM-5 criteria for cannabis use disorder (CUD) or depressive disorders. All participants were required to be Tanner stage ≥4, and female participants were post-menarche.

Participants were excluded if they had: a) a current or past diagnosis of pervasive development disorders, schizophrenia or other psychotic disorders, or substance use disorders other than CUD; b) current use of opioid, sedative, stimulant, and psychedelic substances as indicated on self-reports or urine toxicology tests; c) a full-scale IQ estimate below 80, as determined by the Kaufman Brief Intelligence Test (K-BIT);^29^ d) any acute and/or chronic medical illness, including neurological disorders, as identified on medical history and blood test results; f) any MRI contraindications such as metallic implants, braces, and claustrophobia; and g) a positive pregnancy test.

### 2.3. Cannabis Use Quantification and Dimensional Symptom Measures

Our assessment and quantification of cannabis use were detailed elsewhere^12^ and in our **Supplementary Methods**. Briefly, clinician consensus was reached using combined information from interviews, self-reports, and urine tests to rate each participant based on use frequency and impact on daily function. For our analyses, we coded cannabis use on a 5-level scale: 0 = never used, 1 = tried once, 2 = low use, 3 = moderate use, and 4 = heavy use. Adolescents who did not use cannabis were defined as those who had never used or only tried it once.

Depression severity was measured dimensionally with the clinician-rated Children’s Depression Rating Scale-Revised (CDRS-R)^30^, a validated 17-item instrument with established reliability and validity in adolescents^31^. Items are scored on 1–5 or 1–7 scales, yielding a total score range of 17–113, and specific examples of severity are provided for each item. Higher CDRS-R scores reflect more severe depression.

We also captured related mood and anxiety symptoms using self-reported scales, including the Beck Scale for Suicide Ideation (BSSI)^32^, the Multidimensional Anxiety Scale for Children (MASC)^33^, and the Temporal Experience of Pleasure Scale (TEPS)^34^. The MASC (39 items; range 0–117) indexes anxiety severity, with higher scores indicating greater anxiety. The BSSI (19 items; range 0–38) evaluates suicidal ideation, with 14 follow-up items administered when any of the 5 screening item scores >0. The TEPS measures anticipatory (10 items; range 10–60) and consummatory (8 items; range 8–48) pleasure, with lower scores reflecting higher anhedonia. Anxiety, suicidality, and anhedonia symptom measures served to describe clinical profiles of the cohort and were not included in our primary fMRI analyses.

### 2.4. Neuroimaging Acquisition

Participants completed an MRI clearance procedure to assess for metallic implants or prior exposure to metal. Neuroimaging scans of most participants in this study were performed on a 3T Siemens Skyra scanner (Siemens, Germany) equipped with a 16-channel head coil. A subset of 15 participants was scanned on the same scanner with a 32-channel head receive coil using identical sequences. In all fMRI analyses, coil type was included as a covariate of no interest (binarized as 16- vs. 32-channel).

Imaging protocols aligned with those in the Human Connectome Project (HCP) Lifespan study^35^. Anatomical imaging included a T1-weighted MPRAGE sequence (TR = 2400 ms, TE = 2.06 ms, TI = 1000 ms, flip angle = 8°) with 224 sagittal slices, no interslice gap, a matrix size of 256 × 256, a FOV of 230 × 230 mm^2^, and an isotropic voxel size of 0.9 mm. In addition, a T2-weighted SPACE sequence was acquired (TR = 3200 ms, TE = 565 ms, flip angle = 120°), maintaining the same slice count, matrix, FOV, and voxel resolution as the T1-weighted scan.

Functional imaging was acquired using gradient-recalled echo-planar imaging (EPI). A single resting-state scan (TR = 1000 ms, effective TE = 31.4 ms, flip angle = 60°) lasted 10 minutes and included 600 volumes of 60 slices aligned parallel to the AC-PC plane, with no interslice gaps, a multiband acceleration factor of 5, anterior-to-posterior phase encoding, a matrix size of 98 × 98, an FOV of 228 × 228 mm^2^, and an isotropic voxel size of 2.3 mm. Matched single-band EPI and spin-echo field maps were also acquired for registration and distortion correction. During the resting-state scan, participants were presented with a fixation cross and instructed to keep their eyes open and stay awake. Several task-based fMRI runs unrelated to this study were collected later in the neuroimaging session.

### 2.5. Processing of MRI Data

We visually inspected all data prior to utilizing the HCP Preprocessing Pipelines v3.2^36^. Anatomical preprocessing involved correction for gradient nonlinearity and b_0_ field distortions, followed by alignment to the AC-PC plane, co-registration, skull stripping, and bias field adjustment. Data were then transformed to Montreal Neurological Institute 152 non-linear 6th-generation (MNI152NLin6) space^37^, and cortical segmentation was performed using FreeSurfer. To facilitate surface-based analyses, cortical ribbon extraction was conducted. Functional preprocessing comprised gradient nonlinearity and EPI distortion correction, realignment for motion correction, transformation to MNI space, and intensity normalization. FreeSurfer’s default alignment was used for the initial mapping of functional data onto the cortical ribbon. We then generated participant-level dense timeseries in 32k-CIFTI grayordinate space, merging left and right cortical surfaces with subcortical volumes in MNI space for anatomically accurate representation^36^.

Given the multimodal fMRI design, we applied the multi-run spatial ICA-FIX approach^38–40^ to concatenated resting-state and task runs to identify and remove structured fMRI noise. We conducted mild high-pass filtering (default 2000s cutoff) on concatenated fMRI data and ran ICA using FSL MELODIC on minimally preprocessed whole-brain timeseries in MNI space. Independent components (ICs) were classified as “signal,” “noise,” or “unknown” using the FIX classifier, trained on the default HCP_hp2000.Rdata dataset, which achieves over 97% classification accuracy in locally acquired fMRI datasets. All “signal” and “unknown” ICs were reviewed by experienced neuroimagers in our group and manually reclassified as needed. Subsequently, “noise” ICs were removed from both MNI- and grayordinate-space data using a “soft” regression approach, ensuring that only unique variance associated with noise was eliminated.

Following ICA-FIX denoising, cortical surface data in grayordinate space were aligned using the multimodal surface matching (MSMAll) approach^41^. This multimodal registration method, developed by the HCP^42^, enhances inter-participant correspondence by integrating shared structural and functional features. We applied MSMAll to all resting-state and task runs included in the multi-run ICA-FIX pipeline. Resting-state data were further denoised by regressing out 24 movement parameters (6 affine + temporal derivatives + squares of each) and the five largest principle components associated with white matter and cerebrospinal fluid (CompCor)^43^, identified using CONN Toolbox (RRID:SCR_009550) v17f^44^. Finally, bandpass temporal filtering (0.1 – 0.01Hz) was applied.

### 2.6. fMRI Analyses

Participant-level processed resting-state data were parcellated using a variation of the Cole-Anticevic Brain Network Parcellation (CAB-NP),^45^ which extends the cortical Glasser parcellation^46^ to cover subcortical structures, slightly modified to include somatotopic subdivisions of the somatomotor strip. The resulting 750-node whole-brain data were further restricted to *Reward Expectancy* (114 nodes) and *Reward Attainment* (103 nodes) networks. These network masks were constructed as part of a prior study from nodes with 90^th^ percentile activation strength and their contralateral nodes across the two corresponding contrasts derived from the RFT^20,22,47^ (see details in **Supplementary Methods**).

Consistent with prior work^22^, we used the Brain Connectivity Toolbox v2019–03–03^23^ in MATLAB R2023b (MathWorks, Natick, MA) to estimate three participant-level weighted graph theoretical metrics within each reward network: Strength Centrality (C_Str_), Eigenvector Centrality (C_Eig_), and Local Efficiency (E_Loc_). Specifically, C_Str_ is the sum of all edge weights at a node, reflecting its overall connectivity. Nodes with high C_Str_ maintain strong links with other nodes, indicating robust direct connectivity. C_Eig_ is the eigenvector with the largest eigenvalue for each node and accounts for both a node’s connections and the importance of its neighbors. High C_Eig_ values reflect connections to other highly connected, influential nodes. E_Loc_ describes how efficiently information is exchanged among a node’s immediate neighbors when the node itself is removed. A high E_Loc_ value suggests resilience of local processing despite node disruption. Together, these data-driven metrics captured various influential and communicative properties across nodes within each network^23^.

At the group level, separate generalized linear models (GLM) were performed with each graph theoretical metric (i.e. C_Str_, C_Eig_, E_Loc_) of each network (i.e. *Reward Expectancy* and *Reward Attainment*) serving as the dependent variable. Non-parametric analyses were conducted in FSL PALM^48^ version alpha119 with 10,000 permutations. In all models, independent variables were mean-centered, and correction for family-wise error (FWE) rates across multiple contrasts was applied. Results were considered significant at two-tailed *p_FWE_* < 0.05. To facilitate parallelized group-level analyses, we utilized the *PALM-from-Excel* MATLAB function^49^. All effect sizes were estimated based on *t*-statistics and their associated degrees of freedom (see **Supplementary Methods**). To test our *a priori* hypotheses, we conducted group GLM analyses with cannabis use and depression severity (CDRS-R scores) both included as the responsible variables of interest. This allowed for the assessment of each effect while controlling for the other. In the full sample, the cannabis use variable was binarized to assess for group differences between adolescents who used cannabis and those who did not. Within the group of adolescents who use cannabis, the cannabis use variable included three levels (i.e. low use, moderate use, and heavy use) to examine cannabis use severity. In all models, age, sex, and coil type were included as covariates of no interest. Further, to explore sex differences, we repeated group-level analyses in female and male participants separately, with models adjusted for age and coil type.

To explore the relationship between adolescent cannabis use, depression, and resting-state properties across the whole brain, we repeated the above analyses without participant-level reward network masking. This approach fundamentally changed the computation of C_Str_, C_Eig_, and E_Loc_ metrics within the participant-level timeseries cross-correlation matrix (i.e. 750 × 750 for *Whole Brain* vs. 114 × 114 for *Reward Expectancy* and 103 × 103 for *Reward Attainment*), thereby capturing properties of brain organization beyond reward-related systems.

### 2.7. Other Statistics

In MATLAB R2023b, demographic and clinical profiles of participants were assessed for normality (continuous variables) and expected frequency (categorical variables). Appropriate group comparison and correlation analyses were sequentially conducted, with the significance threshold set at α = 0.05.

## 3. Results

### 3.1. Demographic and Clinical Profiles

The 131 participants (age: 15.3 ± 2.2 years, 65.7% female) included in this study clinically presented across a spectrum of psychiatric illness severity, including those who met DSM criteria for mood, anxiety, and other comorbid psychiatric disorders, those who had subthreshold depression, and those who had no current or past psychiatric diagnoses. Among the 38 participants who used cannabis, 20 reported heavy use (5 met DSM-5 criteria for CUD), 3 reported moderate use, and 15 reported low use. **Table 1** details the demographic and clinical information of study participants.

**Table 1.**
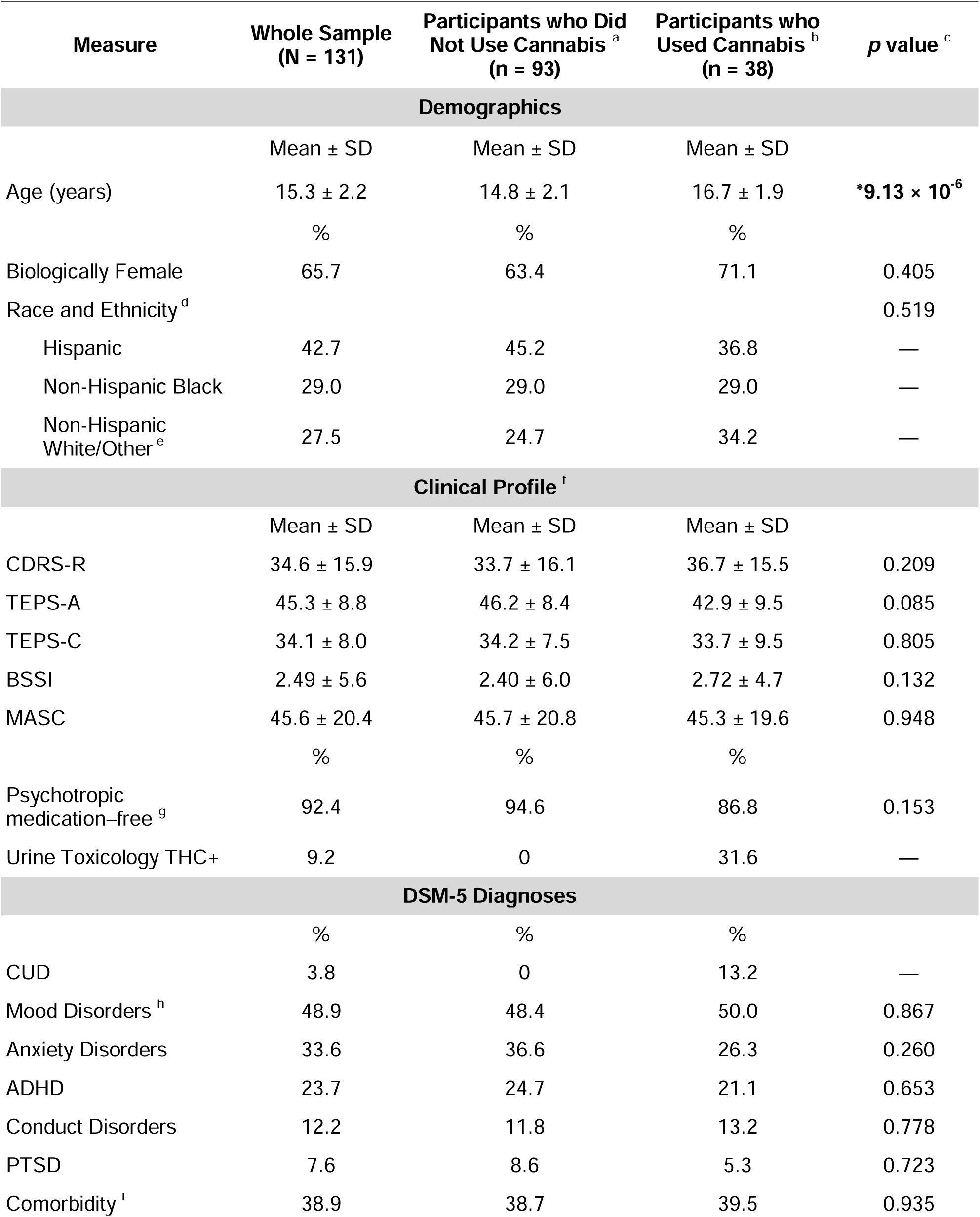
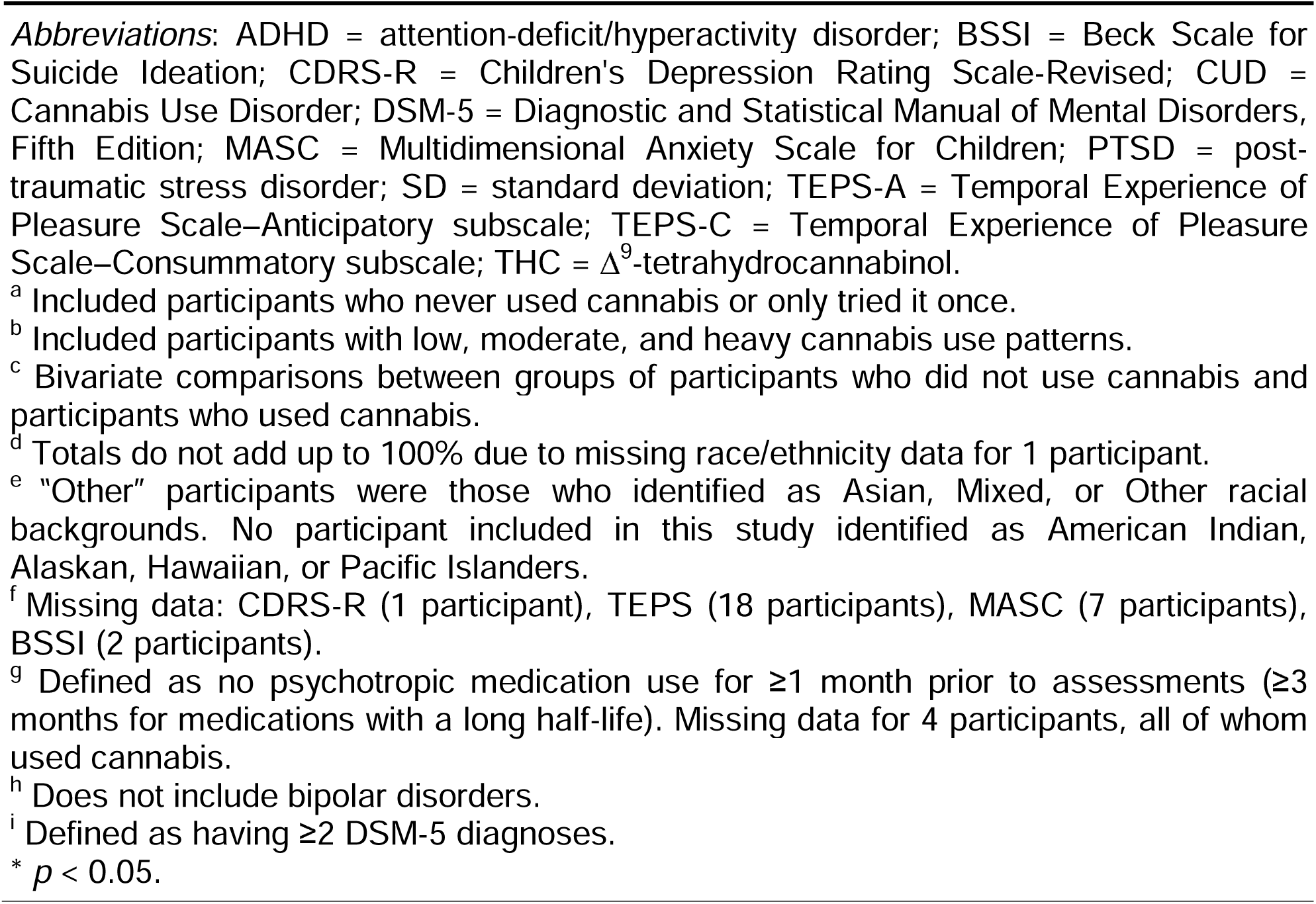
Demographic and clinical characteristics of study participants.

| Measure | Whole Sample<br>(N = 131) | Participants who Did<br>Not Use Cannabis<sup>a</sup><br>(n = 93) | Participants who<br>Used Cannabis<sup>b</sup><br>(n = 38) | p value<sup>c</sup> |
| --- | --- | --- | --- | --- |
| **Demographics** |  |  |  |  |
|  | Mean ± SD | Mean ± SD | Mean ± SD |  |
| Age (years) | 15.3 ± 2.2 | 14.8 ± 2.1 | 16.7 ± 1.9 | **\*9.13 × 10<sup>-6</sup>** |
|  | % | % | % |  |
| Biologically Female | 65.7 | 63.4 | 71.1 | 0.405 |
| Race and Ethnicity<sup>d</sup> |  |  |  | 0.519 |
| Hispanic | 42.7 | 45.2 | 36.8 | — |
| Non-Hispanic Black | 29.0 | 29.0 | 29.0 | — |
| Non-Hispanic<br>White/Other<sup>e</sup> | 27.5 | 24.7 | 34.2 | — |
| **Clinical Profile<sup>f</sup>** |  |  |  |  |
|  | Mean ± SD | Mean ± SD | Mean ± SD |  |
| CDRS-R | 34.6 ± 15.9 | 33.7 ± 16.1 | 36.7 ± 15.5 | 0.209 |
| TEPS-A | 45.3 ± 8.8 | 46.2 ± 8.4 | 42.9 ± 9.5 | 0.085 |
| TEPS-C | 34.1 ± 8.0 | 34.2 ± 7.5 | 33.7 ± 9.5 | 0.805 |
| BSSI | 2.49 ± 5.6 | 2.40 ± 6.0 | 2.72 ± 4.7 | 0.132 |
| MASC | 45.6 ± 20.4 | 45.7 ± 20.8 | 45.3 ± 19.6 | 0.948 |
|  | % | % | % |  |
| Psychotropic<br>medication-free<sup>g</sup> | 92.4 | 94.6 | 86.8 | 0.153 |
| Urine Toxicology THC+ | 9.2 | 0 | 31.6 | — |
| **DSM-5 Diagnoses** |  |  |  |  |
|  | % | % | % |  |
| CUD | 3.8 | 0 | 13.2 | — |
| Mood Disorders<sup>h</sup> | 48.9 | 48.4 | 50.0 | 0.867 |
| Anxiety Disorders | 33.6 | 36.6 | 26.3 | 0.260 |
| ADHD | 23.7 | 24.7 | 21.1 | 0.653 |
| Conduct Disorders | 12.2 | 11.8 | 13.2 | 0.778 |
| PTSD | 7.6 | 8.6 | 5.3 | 0.723 |
| Comorbidity<sup>i</sup> | 38.9 | 38.7 | 39.5 | 0.935 |

*Abbreviations:* ADHD = attention-deficit/hyperactivity disorder; BSSI = Beck Scale for Suicide Ideation; CDRS-R = Children's Depression Rating Scale-Revised; CUD = Cannabis Use Disorder; DSM-5 = Diagnostic and Statistical Manual of Mental Disorders, Fifth Edition; MASC = Multidimensional Anxiety Scale for Children; PTSD = post-traumatic stress disorder; SD = standard deviation; TEPS-A = Temporal Experience of Pleasure Scale—Anticipatory subscale; TEPS-C = Temporal Experience of Pleasure Scale—Consummatory subscale; THC = $\Delta^9$ -tetrahydrocannabinol.
<sup>a</sup> Included participants who never used cannabis or only tried it once.
<sup>b</sup> Included participants with low, moderate, and heavy cannabis use patterns.
<sup>c</sup> Bivariate comparisons between groups of participants who did not use cannabis and participants who used cannabis.
<sup>d</sup> Totals do not add up to 100% due to missing race/ethnicity data for 1 participant.
<sup>e</sup> “Other” participants were those who identified as Asian, Mixed, or Other racial backgrounds. No participant included in this study identified as American Indian, Alaskan, Hawaiian, or Pacific Islanders.
<sup>f</sup> Missing data: CDRS-R (1 participant), TEPS (18 participants), MASC (7 participants), BSSI (2 participants).
<sup>g</sup> Defined as no psychotropic medication use for $\geq 1$ month prior to assessments ( $\geq 3$ months for medications with a long half-life). Missing data for 4 participants, all of whom used cannabis.
<sup>h</sup> Does not include bipolar disorders.

### 3.2. Primary Findings on Cannabis Use and Reward Network Properties

There were no significant group differences in the properties of either reward network between adolescents with vs. without cannabis use. However, in adolescents who used cannabis, heavier cannabis use was correlated with weaker C_Str_ of the right dorsal anterior cingulate cortex (ACC; **Figure 1A**) and weaker E_Loc_ of the left postcentral gyrus (somatosensory cortex; **Figure 1B**) within the *Reward Expectancy* network. These findings were adjusted for depression severity, age, sex, and coil type.

**Figure 1.**
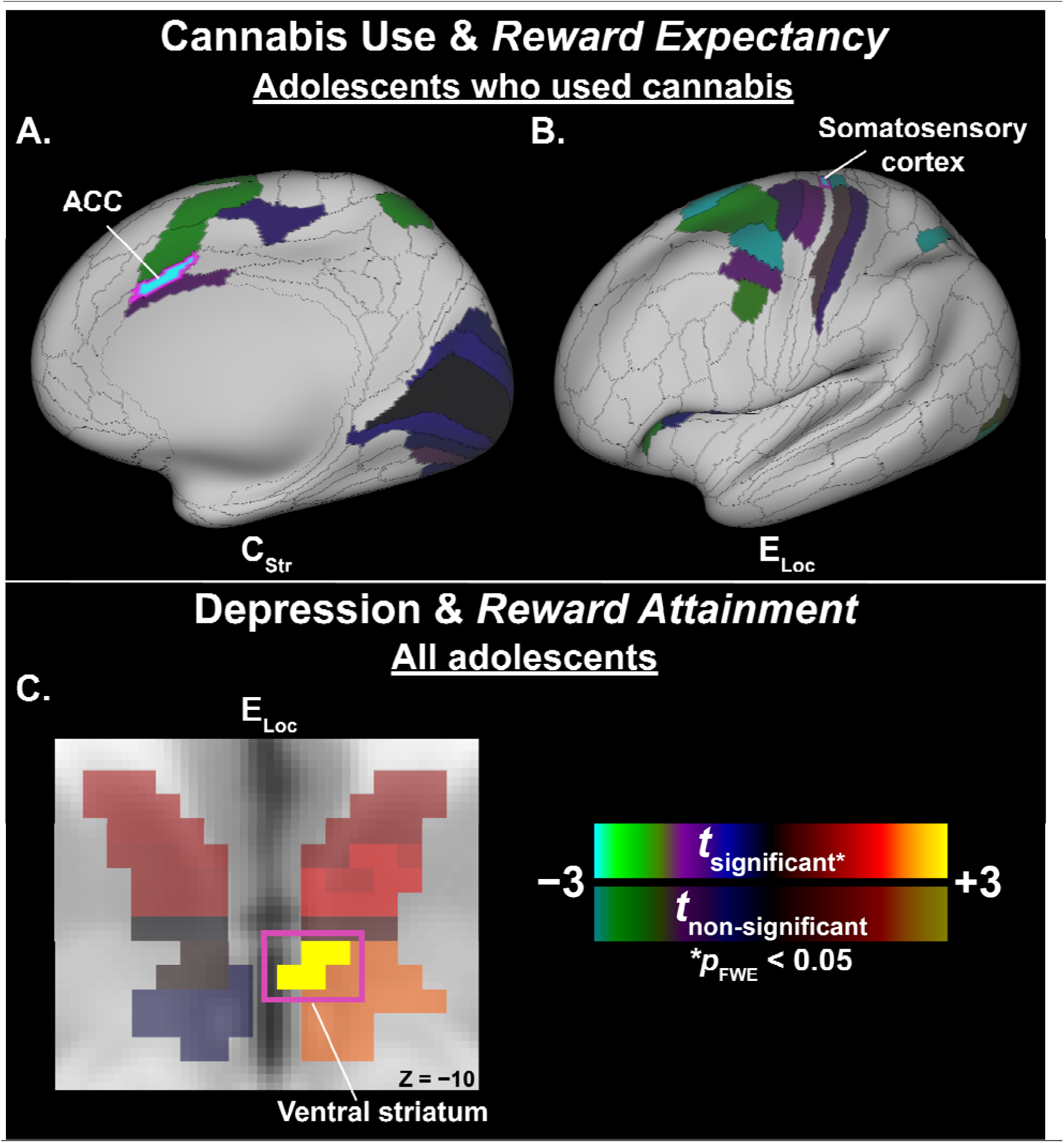
Significant associations between resting-state properties of reward networks and cannabis use and depression severity.

### 3.3. Primary Findings on Depression Severity and Reward Network Properties

In the full sample, depression severity was positively correlated with *Reward Attainment* E_Loc_ of the right ventral striatum (**Figure 1C**), controlling for cannabis use, age, sex, and coil type. When the analysis was restricted to the 38 adolescents who used cannabis, there were no significant associations between depression severity and reward network properties.

**Table 2** lists primary findings with cannabis use and depression.

**Table 2.** Significant alterations in resting-state properties of reward networks in association with adolescent cannabis use and depression.

| Reward Network | Graph Theoretical Metric | Region (Lateralization) | HCP Cortical Label<sup>a</sup> | CAB-NP Label<sup>b</sup> | *t*-statistic<sup>c</sup> | Pearson's *r*<sup>d</sup> | *p*<sub>FWE</sub> |
| --- | --- | --- | --- | --- | --- | --- | --- |
| **Whole Sample (*N* = 131) – Depression Severity<sup>e</sup>** |  |  |  |  |  |  |  |
| Reward Attainment | E<sub>Loc</sub> | Ventral striatum (R) | — | Orbito-Affective-8 | 3.56 | 0.304 | 0.0407 |
| **Adolescents with Cannabis Use (*n* = 38) – Cannabis Use Frequency<sup>f</sup>** |  |  |  |  |  |  |  |
| Reward Expectancy | C<sub>Str</sub> | Anterior cingulate cortex (R) | a24pr | Cingulo-Opercular_9 | −4.08 | −0.585 | 0.0212 |
|  | E<sub>Loc</sub> | Somatosensory cortex (L) | 3a (Trunk)<sup>g</sup> | Somatomotor_30 (Trunk)<sup>g</sup> | −3.64 | −0.541 | 0.0496 |

**Abbreviations:** CAB-NP = Cole-Anticevic Brain-wide Network Partition; FWE = Family-wise error; HCP = Human Connectome Project; L = Left; R = Right.
<sup>a</sup> Labels per the HCP Cortical Atlas, as detailed in Glasser et al. (2016)
<sup>b</sup> Labels per Cole-Anticevic Brain-wide Network Partition v1.0.5 (equivalent labels per HCP S1200 Release cortical parcellation), which comprised 392 cortical nodes derived from HCP parcellation and 358 subcortical nodes from Cole-Anticevic atlas.
<sup>c</sup> Adjusted for age, sex, and head coil type.
<sup>d</sup> Pearson's *r* represents effect size and was computed from *t*-statistic and its corresponding degree of freedom output from FSL PALM.
<sup>e</sup> Depression effect was adjusted for cannabis use effect.
<sup>f</sup> Cannabis use effect was adjusted for depression effect.
<sup>g</sup> Somatotopic subdivision identified per Ely et al.<sup>22</sup> based on secondary data released by the HCP.

### 3.4. Exploratory Sex-Stratified Results

Cannabis use was reported by 31.4% of females (*n* = 27/86) and 24.4% of males (*n* = 11/45), with no significant difference between sexes (*p* = 0.405). Female participants endorsed significantly more severe depression levels than male participants (CDRS-R of females: 36.4 ± 15.9 vs. males: 31.2 ± 15.5; *p* = 0.038).

Male-specific results were significant only in the *Reward Expectancy* analyses. Relative to male youth who did not use cannabis, those who used cannabis showed weaker C_Str_ and C_Eig_ of two nodes in the right thalamus. Among male adolescents who used cannabis, heavier use was associated with weaker C_Str_ of the left thalamus. These findings were controlled for depression severity, age, and coil type.

In female adolescents, only results within the *Reward Attainment* network were significant. Across all female adolescents, depression severity was positively correlated with C_Eig_ of the right ventral diencephalon, after adjusting for cannabis use, age, and coil type. In female adolescents who used cannabis, heavier cannabis use was associated with greater C_Eig_ of the left posterior cingulate cortex (PCC), adjusting for depression severity, age, and coil type.

**Supplementary Figure S1** and **Table S1** detail sex-specific findings.

### 3.5. Exploratory Results from Whole Brain Analyses

Compared to adolescents without cannabis use, those with cannabis use exhibited stronger *Whole Brain* C_Eig_ of the left lateral orbitofrontal cortex (OFC), dorsolateral prefrontal cortex (dlPFC), and posterior inferior frontal junction. Within the group of adolescents who used cannabis, heavier cannabis use correlated with weaker *Whole Brain* C_Str_ and E_Loc_ of the same node within lobule X of the left cerebellum. There was no significant association between depression and *Whole Brain* resting-state properties. Exploratory whole-brain findings are detailed in **Supplementary Figure S2** and **Table S2**.

## 4. Discussion

Bridging across resting-state and task fMRI paradigms, we examined how adolescent cannabis use and depression relate to the intrinsic architecture of reward networks. As hypothesized, cannabis use frequency corresponded to altered properties within the *Reward Expectancy* network, while depression severity was linked to altered properties of the *Reward Attainment* network. Exploratory analyses further revealed: a) male11specific alterations in *Reward Expectancy* and female-specific alterations in *Reward Attainment* network properties; b) relationships between cannabis use and *Whole Brain* resting-state properties. Together, these findings implicate aberrant resting-state organization of reward systems in the co-occurrence of adolescent cannabis use and depression.

### 4.1. Altered Resting-State Properties of Reward Networks in Adolescent Cannabis Use

We observed that lower ACC strength centrality and reduced local efficiency in the somatosensory cortex within the *Reward Expectancy* network scaled with more frequent cannabis use. This is in line with prior longitudinal research showing baseline ACC-OFC hypoconnectivity predicted greater cannabis use over 18 months in adolescents, and that youth with CUD exhibited progressive ACC-prefrontal dysconnectivity relative to controls^50,51^. Although prior resting-state studies of adolescent cannabis use have not implicated somatosensory regions, our finding aligns with recent task-based evidence linking a sensorimotor-prominent cannabis risk network in adolescents to greater addiction severity and poorer treatment outcomes in adults with CUD^52^. While these findings and ours converge on disengagement of the ACC and alteration of somatosensory connectivity in youth with riskier cannabis use behaviors, our focus on the *Reward Expectancy* network adds specificity by localizing these impairments to a reward sub-system. Notably, our *Reward Expectancy* findings parallel our recent observation that adolescent cannabis use is associated with anticipatory anhedonia^12^ and suggest a potential mechanism for such motivational deficits.

In our sex-stratified analyses, results from the male-only sample echoed the primary findings, with cannabis-related alterations restricted to the *Reward Expectancy* network. Interestingly, however, female-only analyses revealed a unique positive correlation between cannabis use frequency and PCC eigenvector centrality within the *Reward Attainment* network. This finding suggests greater influence of the PCC, a key default mode network (DMN) node, on the circuitry underlying the experience of reward in female youth with heavier cannabis use. In contrast to this finding, one previous study found diminished DMN cohesion among female young adults who used cannabis more frequently^53^, while a separate adolescent study reported no significant sex × cannabis use interaction on PCC-based connectivity^54^. These discrepancies may reflect a female-specific developmental trajectory in which cannabinoid effects on PCC connectivity within circuits subserving consummatory reward processing emerge before generalizing to broader DMN dysfunction with continued cannabis use. Given the paucity of research on sex differences in resting-state connectivity and cannabis use, these findings highlight the need to examine how cannabinoids may differentially shape the maturation of neural reward circuitry across sexes.

Our exploratory whole-brain analyses revealed greater global influence of the dlPFC and lateral OFC in adolescents with vs. without cannabis use, along with an association between use frequency and less efficient cerebellar lobule X communication. These results align well with two recent studies documenting greater frontal cortex-based connectivity^55^ and diminished coupling between cerebellar lobule X and the superior frontal gyrus^56^ in youth who used cannabis relative to those who did not. Collectively, these data point to a specific pattern of cannabis-related cerebellar suppression and frontal compensation, reflecting the heightened vulnerability of these CB_1_ receptor-rich regions during adolescence^57,58^.

### 4.2. Altered Resting-State Properties of Reward Networks in Adolescent Depression

Distinct from cannabis use, depression severity was positively correlated with local efficiency of the ventral striatum within the *Reward Attainment* network, reflecting greater striatal network integration in adolescents with worse depression. Adolescent depression has been consistently linked to cortico-striatal dysfunction across a wide range of neuroimaging modalities, including blunted task-evoked striatal responses to rewards^18^ and striatal hyperconnectivity at rest^16,22,59^. Moreover, in a study examining an 11-node reward network in a large youth sample using resting-state fMRI, only elevated baseline striatal strength centrality predicted increased risk of depressive disorder at 3-year follow-up^60^. Consistent with this literature, we found heightened communication efficiency of the striatum with other reward processing regions at rest, potentially as a compensatory reorganization to offset diminished striatal activation during reward attainment.

Our sex-stratified analyses additionally linked depression severity with ventral diencephalon influence within the *Reward Attainment* network in female adolescents. Though exploratory, this finding is consistent with reports of greater vulnerability to depression-related network alterations in female adolescents^61,62^ and further maps this association onto the consummatory reward network. By using task fMRI to functionally localize reward-related networks that can then be examined during rest, our approach underscores the power of cross-modality integration to study reward circuitry with greater specificity in developmental psychopathology.

### 4.3. Clinical Relevance

Our results are highly relevant to clinical settings. Significant disruptions of resting-state networks in co-morbid adolescent cannabis use and depression highlight the importance of screening for both conditions in routine care. These findings also support the utility of existing interventions. In individuals who use cannabis, motivational interviewing and cognitive behavioral therapy (CBT) are particularly effective in identifying motivational deficits and developing personalized, pragmatic strategies to reduce maladaptive coping mechanisms^63,64^. Behavioral activation therapy, a subtype of CBT that increases engagement in rewarding, values-consistent activities, can efficiently enhance enjoyable experiences in adolescents with depression^65^. Our various findings correlating cannabis use frequency and network alterations also support interventions grounded in harm-reduction principles, which prioritize reducing use frequency and promote safer-use education when abstinence is not feasible or desired^66^. Further, our findings dovetail with evidence identifying the ACC as a promising neuromodulation target for addiction in adults^67^. Extending this line of research to youth is critical, as studies on noninvasive brain stimulation for adolescent substance use and comorbid psychopathology remain scarce^68^. Ultimately, mitigating shared reward dysfunction in adolescents with co-occurring cannabis use and depression will require evidence-based treatments to simultaneously target both conditions.

### 4.4. Limitations and Future Directions

There were several limitations in our study. First, the cross-sectional design of this study precludes inferences about causal relationships between detected neural alterations and cannabis use or depression. Future longitudinal studies acquiring data at multiple time points are needed to delineate trajectories of neural and clinical change. Second, the unequal cohort sizes between adolescents with and without cannabis use limited statistical power to detect group differences, with *post hoc* sensitivity analyses indicated 80% power to detect effects of *d* ≥ 0.54 (moderate). For correlation analyses, we achieved 80% power to detect effects of |*r*| ≥ 0.24 (small-to-moderate) in the full sample and |*r*| ≥ 0.42 (moderate-to-large) in adolescents with cannabis use. The proportion of adolescents who used cannabis in this sample (∼29%) was consistent with current epidemiology data^69^, enhancing the generalizability of our findings. While modest, our sample of participants who used cannabis (*n* = 38) was also larger than most resting-state fMRI studies on adolescent cannabis use to date (i.e. typically <30 adolescents who used cannabis)^70^. Nevertheless, future studies should recruit larger cohorts of adolescents who use cannabis, especially those with heavy and disordered use patterns. Third, cannabis use information was primarily self-reported and thus may not be as accurate as quantitative methods based on biological samples, which we plan to incorporate in future work. Quantitative metrics could also facilitate improved dimensionality in correlative analyses. Fourth, our analyses did not adjust for psychosocial confounders such as socio-economic status, family history, and trauma history. Incorporating relevant substance co-use and psychosocial data into future analyses will also clarify factors contributing to brain maturation deviations in this high-risk population.

### 4.5. Conclusions

Our study integrated hypothesis- and data-driven approaches to investigate properties of *Reward Expectancy* and *Reward Attainment* networks during rest in adolescents with co-occurring depression and cannabis use. We identified distinct patterns of reward network disruption associated with each condition and uncovered changes linked to cannabis use that were not previously reported, including sex differences. Taken together, these findings offer new insights into how cannabis use and depression both relate to intrinsic circuitry driving reward function in adolescence, a critical developmental window. Our results challenge the perceived low risks of cannabis use, and we advocate for education efforts and cessation/reduction support for adolescents, especially amid rapidly shifting cannabis policies and increasing product potency. Future research with larger cohorts of adolescents who use cannabis is needed to corroborate and extend these findings.

## Data Availability

All data produced in the present study are available upon reasonable request to the corresponding author.

