## Supplementary Materials for "Altered Reward-Related Resting-State Network Properties in Adolescent Cannabis Use and Depression"

### Abbreviations

ACC: anterior cingulate cortex

AC-PC: anterior commissure–posterior commissure

ADHD: attention-deficit/hyperactivity disorder

BDI: Beck Depression Inventory

BOLD: blood oxygen-level dependent

BSSI: Beck Scale for Suicide Ideation

CAB-NP: Cole-Anticevic Brain Network Partition

$CB_1$: cannabinoid type-1 receptor

CBT: cognitive behavioral therapy

CDRS-R: Children's Depression Rating Scale – Revised

$C_{Eig}$: Eigenvector Centrality

CIFTI: Connectivity Informatics Technology Initiative

CompCor: component correction

$C_{Str}$: Strength Centrality

CUD: cannabis use disorder

dlPFC: dorsolateral prefrontal cortex

DMN: default mode network

DSM: Diagnostic and Statistical Manual of Mental Disorders

$E_{Loc}$: Local Efficiency

EPI: echo-planar imaging

fMRI: functional magnetic resonance imaging

FOV: field of view

FSL: FMRIB Software Library

FWE: family wise error

GLM: generalized linear model

HCP: Human Connectome Project

ICA: independent components analysis

ICA-FIX: ICA-based Xnoiseifier

ICs: independent components

IQ: intelligent quotient

IRB: Institutional Review Boards

K-BIT: Kaufman Brief Intelligence Test

K-SADS-PL: Kiddie Schedule for Affective Disorders and Schizophrenia – Present and Lifetime Version for Children

MASC: Multidimensional Anxiety Scale for Children

MDMA: 3,4-methylenedioxymethamphetamine

MID: monetary incentive delay

MINI: Mini International Neuropsychiatric Interview

MNI: Montreal Neurological Institute

MNI152NLin6: MNI152 Nonlinear 6th generation template

MPRAGE: Magnetization-prepared rapid gradient echo

MRI: magnetic resonance imaging

MSMAll: multimodal surface matching

OFC: orbitofrontal cortex

PALM: Permutation Analysis of Linear Models

PCC: posterior cingulate cortex

PTSD: post-traumatic stress disorder

RFT: Reward Flanker Task

RRID: Research Resource Identifier

SPACE: Sampling Perfection with Application optimized Contrast using different flip angle Evolution

TE: echo time

TEPS: Temporal Experience of Pleasure Scale

TEPS-A: Temporal Experience of Pleasure Scale—Anticipatory subscale

TEPS-C: Temporal Experience of Pleasure Scale—Consummatory subscale

THC: $\Delta^9$-tetrahydrocannabinol

TR: repetition time

### Supplementary Methods

*SM1. Cannabis Use Quantification and Classification*

As detailed in our prior study[1], we combined cannabis use information from K-SADS-PL interviews, urine toxicology results, and self-reports. Study clinicians discussed each case and reached consensus on cannabis use frequency ratings based on the following rationale:

a) Adolescents who reported never using cannabis (coded as 0) or trying it only once (coded as 1) and had negative urine tests were classified as adolescents who did not use cannabis. A single use event was considered a common adolescent experiment, distinct from continued use.
b) Adolescents who reported 2–10 lifetime uses, less than monthly use in the past year without specifying they only tried once, or denied use but tested positive for THC on urine toxicology were classified as those with low use (coded as 2). This level reflected infrequent but ongoing use.
c) Adolescents who reported using cannabis 1–3 times per month over the past year were considered to have "moderate use" (coded as 3). This level indicated substantial but less than weekly use.
d) Adolescents who reported using cannabis weekly to daily and/or met DSM-5 criteria for cannabis use disorder (CUD) were classified as having "heavy use" (coded as 4), as this level of consumption was highly likely to impact daily functioning.

*SM2. Reward Flanker Task and Reward Network Masks*

As described in our prior publication[2], we functionally defined reward network masks by utilizing the fMRI Reward Flanker Task (RFT), designed to elicit activation across multiple phases of reward processing[3,4]. Each RFT trial presented a monetary reward cue, a flanker stimulus, a response period, and reward feedback. Cues comprised certain (50¢, 10¢, or 0¢) and uncertain (?) conditions, with uncertain outcomes equally divided between the three levels of reward. Before scanning, participants learned the task and completed a practice run in a simulated “mock” scanner environment.

Reward network masks were derived from RFT fMRI data collected in a subset of 84 participants from the current study who were previously included in our prior study[2]. These RFT scans were preprocessed using the same HCP-based pipeline described in the main text (see **Methods 2.5**), including multi-run ICAFIX denoising, MSMAll alignment, and parcellation. Data were converted into a “pseudo-NIFTI” format to be compatible with Statistical Parametric Mapping (SPM) v12 for participant-level analyses. BOLD signals were modeled with a generalized linear model including 11 regressors (4 cue types, 3 correct feedback types following certain cues, 3 correct feedback types following uncertain cues, and an error regressor). Activation contrasts were defined as:

1. *Reward Expectancy* = (50¢ + 10¢ reward cues) – (0¢ non-reward cues)
2. *Reward Attainment* = (50¢ + 10¢ reward feedback) – (0¢ non-reward feedback)

Prior to group-level analysis, participant-level contrast maps were reformatted into CIFTI. Group-level activation for each reward phase was estimated on parcellated RFT data using FSL PALM[5] v111-alpha. For each network, the top 10th percentile of activated parcels (nodes) was retained. To avoid artificial lateralization from thresholding, corresponding contralateral nodes were also included, yielding symmetrical network masks. The final *Reward Expectancy* and *Reward Attainment* network masks included 114 and 103 nodes, respectively.

*SM3. Computations of Effect Sizes for Significant fMRI Results*

All effect sizes were computed from the permutation-derived *t*-statistic output by FSL PALM. Effect sizes for significant correlations (cannabis use frequency and depression severity) were reported as Pearson’s *r*, which was derived from the *t*-statistic and its corresponding degrees of freedom using the formula[6,7]: $r = \sqrt{\frac{t^2}{t^2+df}}$. For significant two-sample group comparisons (adolescents with vs. without cannabis use), we first converted the *t*-statistic to Cohen's *d* using the formula[6]: $d = t\sqrt{\frac{1}{n_1} + \frac{1}{n_2}}$. We then applied Hedges' small-sample bias correction to obtain *g* as follows[8]: $g = d(1 - \frac{3}{4(n_1+n_2)-9})$. Reporting group comparison effect sizes as Hedges' *g* is more appropriate than Cohen's *d*, given the relatively modest size of the group of adolescents who used cannabis compared to the group of adolescents who did not use cannabis[8].

*SM4.* Post Hoc *Sensitivity Analyses*

*Post hoc* sensitivity analyses were conducted using G*Power v3.1.9.7 software[9] for group comparisons (38 adolescents who used cannabis vs. 93 adolescents who did not use cannabis) and correlations across the full sample ($N$ = 131) and within the group of adolescents who used cannabis ($n$ = 38).

### Supplementary Results

*SR1. Participants*

Data from 79 individuals included in our prior publication[2] were incorporated in this study.

*SR2. Exploratory Findings on Sex Differences*

**Supplementary Figure 1.** Sex differences between relationships of resting-state properties of reward networks and cannabis use and depression severity.

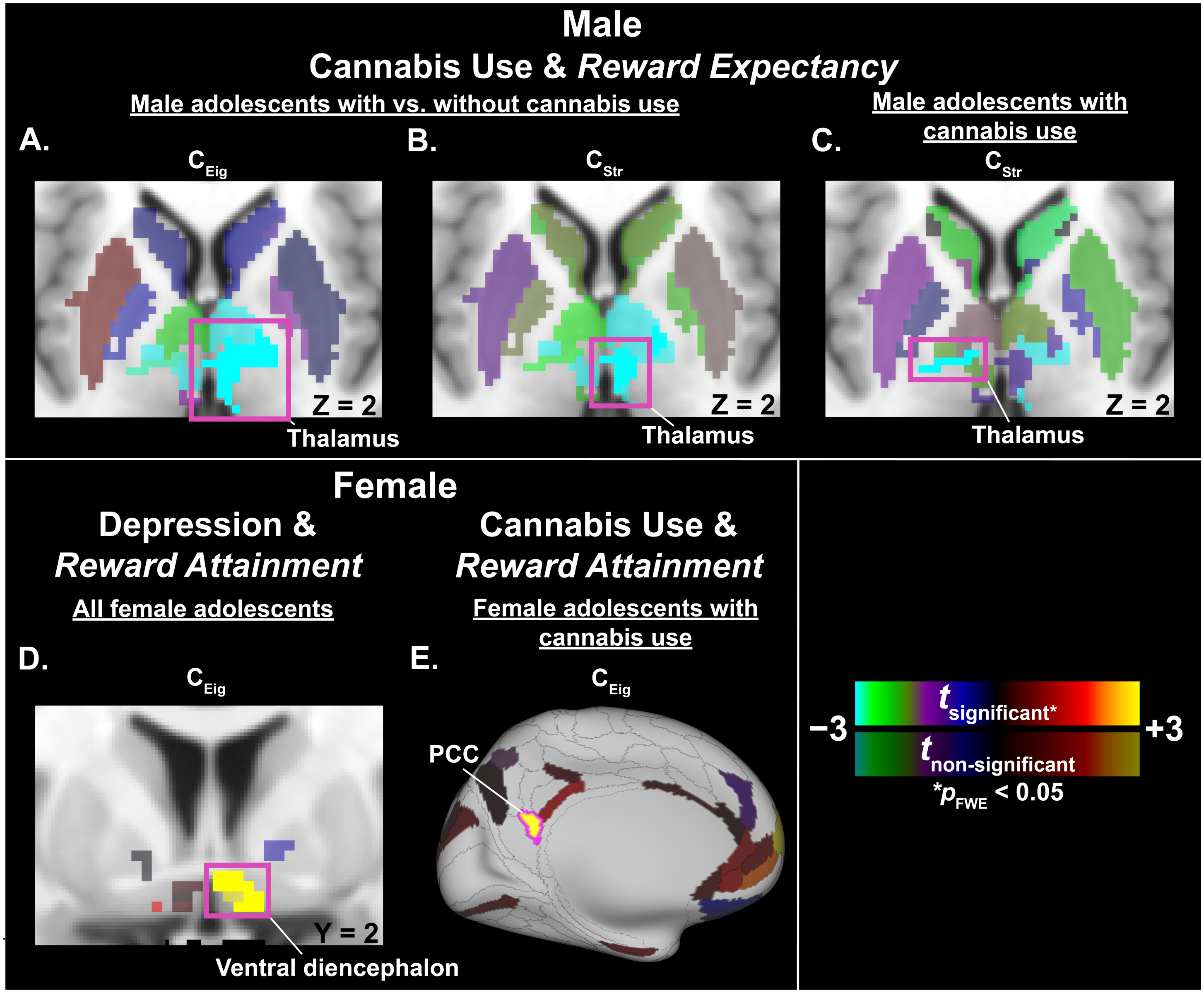

*Abbreviation:* $C_{Eig}$ = eigenvector centrality; $C_{Str}$ = strength centrality; FWE = family-wise error; PCC = posterior cingulate cortex.

Relative to male adolescents without cannabis use, those who used cannabis exhibited weaker **(A)** eigenvector centrality and **(B)** strength centrality of the right thalamus within the *Reward Expectancy* network. In male adolescents who used cannabis, **(C)** heavier cannabis use was negatively correlated with strength centrality of the right thalamus within the *Reward Expectancy* network. In all female adolescents, **(D)** depression severity correlated with stronger eigenvector centrality of the left ventral diencephalon within the *Reward Attainment* network. In female adolescents who used cannabis, **(E)** heavier cannabis use positively correlated with eigenvector centrality of the left PCC within the *Reward Expectancy* network. Results are adjusted for depression severity, age, and head coil type, and displayed on axial (**A**, **B**, **C**) and coronal (**D**) slices through subcortical MNI space, as well as the 'very inflated' left cortical surface (**E**) in Connectome Workbench v2.0.0. Significant parcels (two-tailed $p_{FWE} < 0.05$) are outlined in fuchsia and labeled. Non-significant *t*-statistics are shown at 50% opacity relative to significant *t*-statistics.

**Supplementary Table S1.** Significant sex-stratified results

| Reward Network | Graph Theoretical Metric | Region (Lateralization) | HCP Cortical Label [a] | CAB-NP Label [b] | *t*-statistic [c] | Effect Size [d] | $p_{FWE}$ |
|---|---|---|---|---|---|---|---|
| ***All Female Adolescents** (n = 86) – **Depression Severity** [e]* | | | | | | | |
| *Reward Attainment* | $C_{Eig}$ | Ventral diencephalon (R) | — | Default-20 | 3.79 | 0.388 | 0.0284 |
| ***Female Adolescents with Cannabis Use** (n = 27) – **Cannabis Use Frequency** [e]* | | | | | | | |
| *Reward Attainment* | $C_{Eig}$ | Posterior cingulate cortex (L) | v23ab | Default_40 | 4.06 | 0.655 | 0.0452 |
| ***Male Adolescents with** (n = 11) **vs. without** (n = 34) **Cannabis Use*** | | | | | | | |
| *Reward Expectancy* | $C_{Eig}$ | Thalamus (R) | — | Cingulo-Opercular-38 | −4.27 | −1.46 | 0.0135 |
| | | Thalamus (R) | — | Visual-63 | −3.97 | −1.35 | 0.0311 |
| | $C_{Str}$ | Thalamus (R) | — | Cingulo-Opercular-38 | −3.63 | −1.24 | 0.0434 |
| ***Male Adolescents with Cannabis Use** (n = 11) – **Cannabis Use Frequency** [e]* | | | | | | | |
| *Reward Expectancy* | $C_{Str}$ | Thalamus (L) | — | Visual-59 | −9.96 | −0.971 | $7.10 \times 10^{-3}$ |

*Abbreviations*: CAB-NP = Cole-Anticevic Brain-wide Network Partition; FWE = Family-wise error; HCP = Human Connectome Project; L = Left; R = Right.

[a] Labels per the HCP Cortical Atlas, as detailed in Glasser et al. (2016)[10]

[b] Labels per Cole-Anticevic Brain-wide Network Partition v1.0.5 (equivalent labels per HCP S1200 Release cortical parcellation), which comprised 392 cortical nodes derived from HCP parcellation and 358 subcortical nodes from Cole-Anticevic atlas.

[c] Adjusted for age and head coil type.

[d] Reported as Hedges' *g* for group comparisons and Pearson's *r* for correlations.

[e] Cannabis use effect was adjusted for depression effect.

[f] Depression effect was adjusted for cannabis use effect.

*SR3. Exploratory Whole-Brain Findings*

**Supplementary Figure 2.** Significant associations between whole-brain resting-state properties and cannabis use.

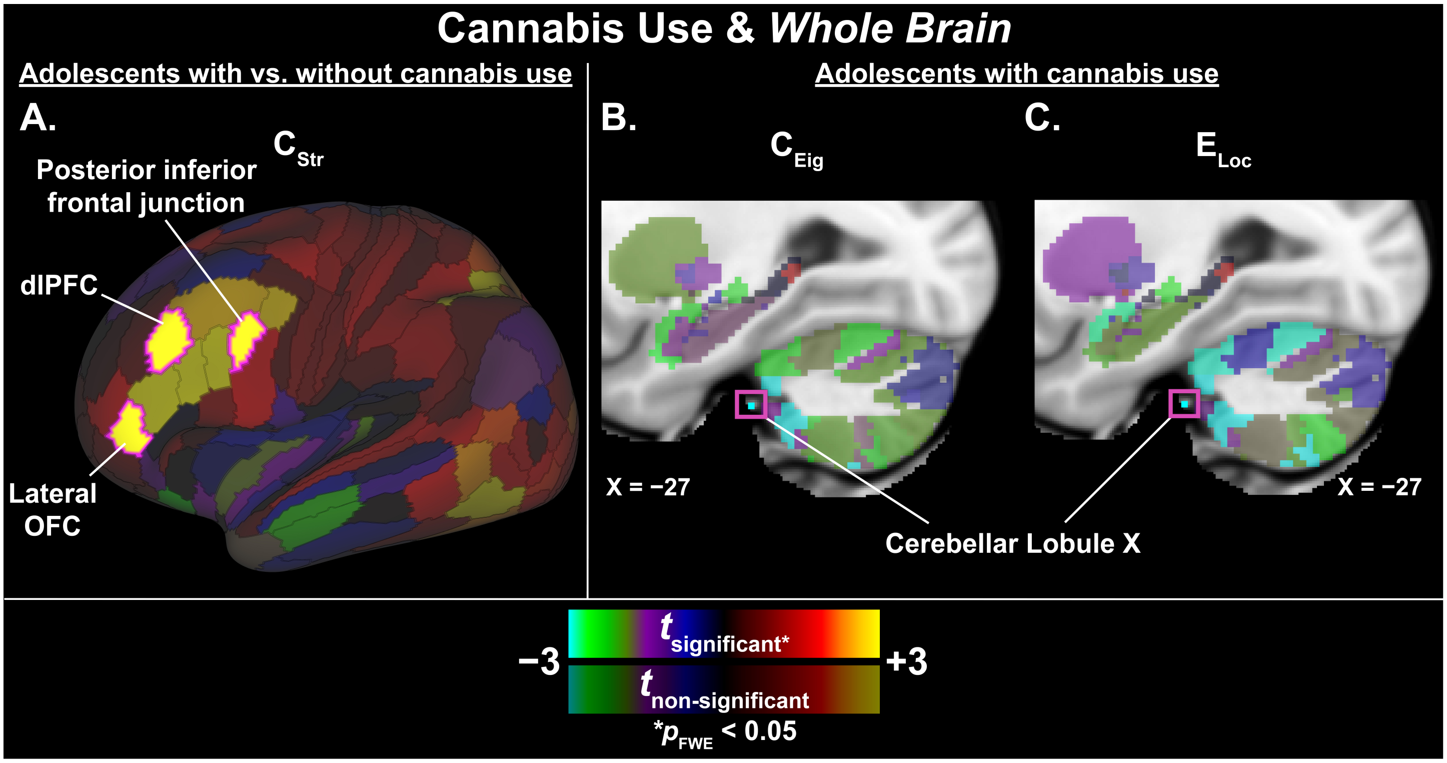


*Abbreviation:* $C_{Eig}$ = eigenvector centrality; $C_{Str}$ = strength centrality; dlPFC = dorsolateral prefrontal cortex; $E_{Loc}$ = local efficiency; FWE = family-wise error; OFC = orbitofrontal cortex.

Relative to adolescents without cannabis use, those who used cannabis exhibited **(A)** stronger strength centrality of left frontal regions. In adolescents who used cannabis, significant correlations were observed between heavier cannabis use and weaker **(B)** eigenvector centrality and **(C)** local efficiency of the same node within lobule X of the left cerebellum. Results are adjusted for depression severity, age, sex, and head coil type, and displayed on the 'very inflated' left cortical surface (**A**) and a sagittal slice through subcortical MNI space (**B**, **C**) in Connectome Workbench v2.0.0. Significant parcels (two-tailed $p_{FWE} < 0.05$) are outlined in fuchsia and labeled. Non-significant *t*-statistics are shown at 50% opacity relative to significant *t*-statistics.

**Supplementary Table S2.** Significant whole-brain results

| Network | Graph Theoretical Metric | Region (Lateralization) | HCP Cortical Label [a] | CAB-NP Label [b] | *t*-statistic [c] | Effect Size [d] | $p_{FWE}$ |
|---|---|---|---|---|---|---|---|
| ***Adolescents with** (n = 38) **vs. without** (n = 93) **Cannabis Use** [e]* | | | | | | | |
| *Whole Brain* | $C_{Eig}$ | Lateral orbitofrontal cortex (L) | p47r | Frontoparietal_49 | 4.93 | 0.944 | $2.20 \times 10^{-3}$ |
| | | Dorsolateral prefrontal cortex (L) | p9-46v | Frontoparietal_36 | 4.26 | 0.816 | 0.0231 |
| | | Posterior inferior frontal junction (L) | IFJp | Frontoparietal_34 | 5.19 | 0.994 | $1.00 \times 10^{-3}$ |
| ***Adolescents with Cannabis Use** (n = 38) – **Cannabis Use Frequency** [e]* | | | | | | | |
| *Whole Brain* | $C_{Str}$ | Cerebellar lobule X (L) | — | Frontoparietal-15 | −4.49 | −0.621 | 0.0346 |
| | $E_{Loc}$ | Cerebellar lobule X (L) | — | Frontoparietal-15 | −4.44 | −0.618 | 0.0304 |

*Abbreviations*: CAB-NP = Cole-Anticevic Brain-wide Network Partition; FWE = Family-wise error; HCP = Human Connectome Project; L = Left.

[a] Labels per the HCP Cortical Atlas, as detailed in Glasser et al. (2016)[10]

[b] Labels per Cole-Anticevic Brain-wide Network Partition v1.0.5 (equivalent labels per HCP S1200 Release cortical parcellation), which comprised 392 cortical nodes derived from HCP parcellation and 358 subcortical nodes from Cole-Anticevic atlas.

[c] Adjusted for age, sex, and head coil type.

[d] Reported as Hedges' *g* for group comparisons and Pearson's *r* for correlations.

[e] Cannabis use effect was adjusted for depression effect.
